# A Reproducible Health Informatics Pipeline for Simulating and Integrating Early-Phase Oncology Clinical, Biomarker, and Pharmacokinetic Data for Exploratory Decision-Support Analytics

**DOI:** 10.64898/2026.03.27.26349538

**Authors:** Mark I.R. Petalcorin

**Affiliations:** aAidea, London, United Kingdom

**Keywords:** Health Informatics, oncology, phase I trial, biomarkers, pharmacokinetics, reproducibility, simulation, machine learning, translational analytics, clinical data integration

## Abstract

**Background:** Early-phase oncology development increasingly depends on integrated interpretation of clinical outcomes, translational biomarkers, and pharmacokinetic exposure rather than toxicity alone. This shift has created a need for reproducible analytical workflows that can combine heterogeneous trial data into traceable, analysis-ready outputs suitable for exploratory review and early decision support.

**Objective:** To develop a reproducible Python-based workflow that simulates a plausible early-phase oncology study, integrates clinical, biomarker, and pharmacokinetic data, and generates analysis-ready datasets, visual summaries, and exploratory predictive models relevant to early development analytics.

**Methods:** A workflow was constructed to simulate an early-phase oncology cohort of 120 patients distributed across multiple dose levels. Three synthetic raw data sources were generated, including patient-level clinical data, baseline biomarker data, and longitudinal pharmacokinetic profiles. These sources were merged into a single analysis-ready dataset containing derived variables such as tumor percent change from baseline, clinical-benefit status, exposure summaries, adverse-event indicators, and survival outcomes. The workflow produced structured tables, patient listings, waterfall plots, Kaplan-Meier-style survival curves, biomarker-response visualizations, pharmacokinetic profile plots, and exploratory machine-learning outputs.

**Results:** The final integrated dataset contained 120 patients and 30 variables. Median survival across the simulated cohort was 243.8 days, and higher dose groups showed improved median survival and greater clinical benefit relative to the low-dose group. Clinical benefit increased from 8.6% in the low-dose group to 29.0% in the medium-dose group and 45.2% in the high-dose group. Higher baseline LDH, CRP, and ctDNA fraction tracked with less favorable tumor-response trajectories, whereas higher exposure, reflected by AUC and Cmax, associated with improved disease control. Pharmacokinetic profiles showed clear dose-dependent separation. Grade 3 or higher adverse-event rates remained within a plausible exploratory range across dose groups. A random-forest model for clinical benefit achieved an exploratory ROC AUC of 0.845, while a logistic-regression model for strict responder status could not be fit because no simulated patient met the prespecified objective response threshold.

**Conclusions:** This proof-of-concept demonstrates that a transparent Python workflow can generate a coherent early-phase oncology analytical ecosystem from synthetic inputs. The workflow supports integration of heterogeneous data streams, derivation of analysis-ready variables, production of interpretable outputs, and exploratory modeling in a reproducible framework. Although the simulated responder prevalence was too low to support objective response modeling, this limitation itself highlights the importance of simulation calibration for downstream analytical validity. The framework provides a practical Health Informatics demonstration of how early oncology trial data can be structured and analyzed for exploratory translational decision support.

## Introduction

Early-phase oncology development has changed substantially over the last two decades. Historically, many phase I studies were designed primarily to identify a maximum tolerated dose, often using rule-based escalation schemes in which toxicity served as the dominant decision variable. In contemporary oncology, that framework has broadened considerably. Investigators now seek not only to determine whether a drug can be administered safely, but also to understand whether a given dose produces a biologically meaningful exposure, whether it modulates the intended target, whether early signs of antitumor activity emerge, and whether those effects occur within an acceptable safety window. This shift from simple dose finding toward dose optimization has been driven by the rise of targeted therapies, biologics, and immuno-oncology agents, for which the most informative dose is not always the highest tolerable one, but rather the dose that best balances exposure, pharmacodynamic effect, antitumor activity, and tolerability (Gibbs et al., 2010; Wang et al., 2017; Goldstein & Peters, 2021; Solans et al., 2020).

That evolution has created growing demand for integrated analytical workflows capable of linking multiple types of trial data in a reproducible way. In practice, early-development teams rarely review a single endpoint in isolation. Instead, they must synthesize baseline clinical characteristics, laboratory and circulating biomarkers, serial pharmacokinetic measurements, adverse-event summaries, and radiographic or categorical tumor outcomes into a traceable analytical structure. These integrated datasets support much more than descriptive review. They help teams explore exposure-response relationships, generate translational hypotheses, contextualize early efficacy signals, and inform decisions about expansion cohorts, biomarker strategy, and later-stage study design. Phase I oncology studies that incorporate both PK and PD components have illustrated the value of this integrated view, especially when early biological activity is modest and must be interpreted in combination with exposure and safety data rather than through response rate alone (Gibbs et al., 2010; Solans et al., 2020; Mahadevan et al., 2011).

Within solid-tumor oncology, radiographic interpretation still relies heavily on RECIST-based concepts. Percent change in tumor burden remains one of the most familiar practical summaries of antitumor effect in early clinical studies, even when investigators recognize that it captures only one dimension of treatment activity. RECIST 1.1 and its subsequent clarifications provide widely accepted thresholds for response and progression, and these thresholds continue to shape how investigators visualize and communicate tumor outcomes. At the same time, survival-type endpoints remain central because they provide important context for dose groups and biomarker-defined subgroups, particularly when formal objective responses are rare. Even in exploratory settings, time-to-event analyses help reveal whether disease control, treatment exposure, or biomarker burden associate with longer or shorter clinical trajectories. However, survival curves must be interpreted with awareness of censoring, competing risks, and other structural limitations that can influence apparent group differences (Eisenhauer et al., 2009; Schwartz et al., 2016; Fleming & Lin, 2000; D’Arrigo et al., 2021; Gomes et al., 2024).

The increasing role of translational biomarkers has further transformed early-phase oncology. Several circulating or routine laboratory variables are attractive precisely because they can be measured repeatedly and with relatively low burden, allowing investigators to follow both disease biology and host response over time. Circulating tumor DNA, or ctDNA, has emerged as one of the most promising of these biomarkers. It can provide noninvasive information about molecular disease burden, treatment response, emergence of resistance, and trial enrichment strategies, making it especially relevant in modern early-phase programs where serial tissue biopsies are often limited or impractical (Parisi et al., 2023; Tan et al., 2026). At the same time, more conventional markers remain clinically valuable. Lactate dehydrogenase, LDH, has repeatedly shown prognostic significance across multiple solid tumors and is often interpreted as a marker of aggressive tumor biology or metabolic stress, whereas C-reactive protein, CRP, is a broad inflammatory marker that has been associated with poorer prognosis, recurrence, and treatment response in adults with solid malignancies (Petrelli et al., 2015; Shrotriya et al., 2015). ECOG performance status also remains one of the most robust clinical summary variables in oncology, with systematic reviews showing that poorer performance status consistently associates with worse outcomes across multiple disease contexts (Assayag et al., 2023; Dall’Olio et al., 2020).

As these data streams grow richer, interest in predictive analytics has expanded in parallel. Regression models, tree-based methods, and other machine-learning approaches are increasingly used to explore response, prognosis, patient stratification, and biomarker-outcome relationships. Yet the clinical-prediction literature has repeatedly warned that algorithmic sophistication alone does not guarantee practical value. Across medical prediction studies, machine-learning approaches have often failed to show consistent performance gains over logistic regression, especially when comparisons are poorly designed or reported. Equally important, the utility of any predictive model depends on transparent reporting, realistic validation, and clarity about intended use. In oncology specifically, recent reviews show that although many machine-learning models have been published, far fewer have undergone robust external validation or demonstrated clear clinical usefulness in real decision-making settings (Christodoulou et al., 2019; Collins et al., 2015; Collins et al., 2024; Santos & Amorim-Lopes, 2025).

For these reasons, a simulated proof-of-concept workflow is useful as both a methodological and pedagogic tool. It allows investigators to demonstrate how heterogeneous early-phase data can be structured, merged, derived, summarized, visualized, and modeled without relying on real patient records. Such an approach is especially valuable for showcasing data flow, endpoint definition, exposure summarization, exploratory modeling logic, and output generation in a transparent environment. It also creates a sandbox in which one can test whether the analytical workflow itself is coherent before moving to real-world data, which may be constrained by privacy, missingness, operational noise, or inconsistent collection practices. The aim of the present work is therefore to build a reproducible Python workflow that simulates a plausible early-phase oncology study, integrates clinical, biomarker, and pharmacokinetic data into an analysis-ready dataset, and generates outputs representative of exploratory decision-support analytics used in modern oncology development.

## Methods

### Study concept and computational design

A script-based workflow was constructed to simulate an early-phase oncology dataset and to reproduce a practical sequence of early development analyses. The workflow was organized to mirror a modular programming repository, with components corresponding to raw-data simulation, dataset integration, table generation, visualization, and exploratory modeling. The workflow generated three raw sources, clinical data, biomarker data, and pharmacokinetic data, followed by a merged analysis_dataset.csv, summary tables, figures, and model outputs. All analyses were executed in Python with fixed random seeding for reproducibility. The computational environment used common scientific and machine-learning libraries, including NumPy, pandas, Matplotlib, and scikit-learn. The overall design followed the general principle that clinical prediction and exploratory analytics pipelines should be transparent, modular, and reproducible, even when the goal is methodological demonstration rather than formal clinical deployment (Collins et al., 2015; Collins et al., 2024).

### Simulated population and dose structure

The synthetic trial included 120 simulated patients distributed across four nominal dose levels of 25, 50, 100, and 200 mg. For interpretability, these doses were collapsed into three descriptive dose groups: low, consisting of 25 and 50 mg, medium, consisting of 100 mg, and high, consisting of 200 mg. Baseline clinical variables included patient identifier, age, sex, ECOG performance status, dose level, dose group, and baseline tumor burden. Age was simulated between 32 and 82 years. ECOG status was sampled from values 0, 1, and 2 with probabilities favoring ECOG 0 to 1, consistent with the fact that early-phase oncology studies commonly enroll patients with preserved performance status (Assayag et al., 2023; Dall’Olio et al., 2020). Baseline tumor burden was sampled from a normal distribution centered near 95 mm and truncated to a plausible range for measurable disease. The intent was not to emulate any specific tumor type, but to provide a continuous disease-burden variable that could feed subsequent size-based and time-to-event simulations in a RECIST-like framework (Eisenhauer et al., 2009; Schwartz et al., 2016).

### Biomarker simulation

For each patient, a biomarker table was simulated containing mutation status, LDH, CRP, glucose, lactate, ctDNA fraction, and a composite biomarker-response score. Mutation status was binary, with mutant status assigned to approximately 40% of patients. LDH and CRP were sampled from log-normal distributions to generate positively skewed values, reflecting the fact that these biomarkers often show right-skewed distributions in oncology cohorts. ctDNA fraction was simulated from a beta-distributed variable scaled into a low-fraction range. This design reflected the practical role of ctDNA as a circulating biomarker for real-time assessment of tumor dynamics, while LDH and CRP were incorporated because of their recognized associations with tumor burden, inflammation, and adverse prognosis (Tan et al., 2026; Parisi et al., 2023; Petrelli et al., 2015; Shrotriya et al., 2015). A composite biomarker-response score was then generated as a weighted standardized combination of LDH, CRP, and ctDNA plus Gaussian noise. This score was not used as a regulatory endpoint. Instead, it acted as a latent translational variable to make the simulated dataset internally coherent, with worse biomarker burden tending to align with worse downstream tumor and survival outcomes.

### Clinical endpoint simulation

Clinical outcomes were generated by combining dose effect, biomarker penalties, ECOG penalty, mutation-associated benefit, and random noise. The dose effect was specified such that higher doses moved simulated tumor change in a more favorable direction. In contrast, higher LDH, higher CRP, higher ctDNA fraction, and worse ECOG status shifted tumor change toward progression. Mutant status conferred a modest favorable effect. Week 6 tumor size was then calculated by applying the simulated progression ratio to baseline tumor size. Tumor percent change from baseline was derived as:

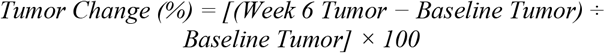

Best overall response was assigned using RECIST-like categories based on the simulated percent change. Patients with tumor change less than or equal to −30% were labeled partial response, values between −30% and +20% were labeled stable disease, and values of +20% or greater were labeled progressive disease. These thresholds were chosen because they are consistent with the size-based response logic that underlies RECIST 1.1, while recognizing that this project does not implement full formal RECIST adjudication (Eisenhauer et al., 2009; Schwartz et al., 2016). A binary Responder endpoint was defined as tumor change less than or equal to −30%. A broader binary Clinical_Benefit endpoint was defined as either responder status or best overall response in the categories stable disease, partial response, or complete response. This broader endpoint was included because in many early development settings, disease stabilization and early biological control can be informative even when objective response is uncommon, particularly for targeted or cytostatic agents (Goldstein et al., 2021; Mahadevan et al., 2011).

### Adverse event simulation

Adverse-event severity was generated as a dose- and risk-dependent process. A severe adverse event probability was computed from LDH, dose, and ECOG. Using that probability, each patient was assigned an adverse event grade from 1 to 4. Binary variables for grade 3+ adverse events, serious adverse events, and treatment discontinuation were then derived. This approach was designed to reproduce a clinically recognizable early-phase structure in which toxicity tends to increase with dose and patient frailty, although not perfectly monotonically in a small simulated cohort. The logic reflects the central role of safety profiling in phase I oncology studies, even as modern studies increasingly combine safety with exposure and biomarker interpretation (Gibbs et al., 2010; Mahadevan et al., 2011).

### Survival simulation

Survival time and event status were generated using a risk score combining LDH, CRP, ctDNA fraction, ECOG, dose level, and best overall response class. Higher biomarker burden and poorer response increased hazard, whereas higher dose and favorable response lowered hazard. Simulated survival time was truncated to a plausible range and event probability was generated through a logistic transform of the risk score. This yielded a dataset suitable for Kaplan-Meier-style descriptive analysis by dose group and by biomarker-defined subgroup. The present project intentionally used a simple product-limit style estimator for visual survival display, rather than a more elaborate semiparametric or competing-risks framework. Kaplan-Meier curves remain the most familiar and widely used descriptive survival display in medical research, but interpretation must remain cautious where competing risks or nonproportional hazards are relevant (Fleming & Lin, 2000; D’Arrigo et al., 2021; Gomes et al., 2024).

### Pharmacokinetic simulation and exposure summaries

Longitudinal PK measurements were simulated at predefined times of 0, 1, 2, 4, 8, 24, 48, and 72 hours. Each patient’s profile was generated to show increasing concentration with dose, an early peak, and subsequent decay. Patient-level PK summaries included area under the concentration-time curve from 0 to 72 hours, written as AUC_0_–_72_, maximum concentration, written as Cmax, and time of maximum concentration, written as Tmax_hr. AUC was calculated using the trapezoidal rule after ordering time points. This design was chosen because exposure metrics such as AUC and Cmax are central to evaluating exposure-response and exposure-toxicity relationships in oncology drug development, especially when doses with similar nominal intensity may still produce substantial interpatient variability in systemic exposure (Solans et al., 2020; Wang et al., 2017; Castellano et al., 2020).

### Analysis-ready dataset construction

The core processed file, analysis_dataset.csv, merged clinical outcomes, biomarker variables, and PK summaries by patient identifier. Derived variables included tumor percent change from baseline, responder status, clinical benefit, high-versus-low biomarker indicators, and AE grade 3+ status. This step was conceptually analogous to building an analysis-ready dataset for integrated exploratory review. In addition, summary_table.csv was generated by dose group and included sample size, mean age, percent female, mean baseline tumor size, responder rate, median survival, mean LDH, mean CRP, grade 3+ AE rate, serious AE rate, and discontinuation rate. A separate patient_listing.csv was produced for patient-level review.

### Visualizations

The workflow generated standard exploratory figures: Kaplan-Meier-style survival curves were drawn by dose group; a bar chart summarized the rate of grade 3+ adverse events by dose group; a waterfall plot displayed patient-level tumor percent change sorted from best to worst response, with dashed reference lines at −30% and +20% corresponding to RECIST-like response thresholds; a scatter plot examined the relationship between baseline LDH and tumor percent change; and, mean PK profiles were shown by dose group. These plots were selected because they are familiar to oncology development teams and serve distinct interpretive functions. Waterfall plots provide intuitive visualization of heterogeneity in tumor shrinkage, while Kaplan-Meier curves remain the default descriptive display for time-to-event outcomes in trials (Eisenhauer et al., 2009; Gomes et al., 2024). The early-phase clinical biomarker pipeline was organized as a reproducible conceptual workflow (Figure 1) that begins with simulated raw clinical, biomarker, and pharmacokinetic data inputs and proceeds through sequential stages of dataset construction, visualization, statistical modeling, and generation of review-ready outputs. This framework was designed to mirror the structure of a practical early-phase oncology analytical pipeline, in which heterogeneous data sources are integrated into an analysis-ready dataset and then used to produce exploratory figures, summary tables, and predictive modeling outputs for downstream interpretation and decision support.

**Figure 1.**
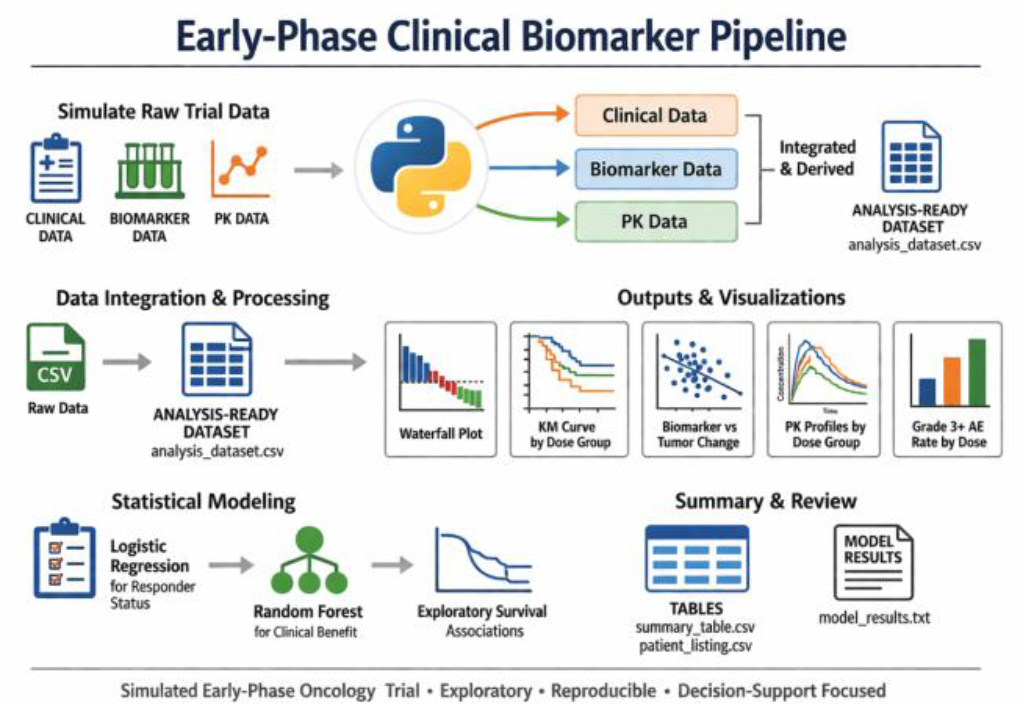
Schematic overview of the early-phase clinical biomarker pipeline. Conceptual workflow diagram illustrating the structure of the reproducible early-phase oncology analysis pipeline, from simulated raw clinical, biomarker, and pharmacokinetic inputs through dataset construction, visualization, statistical modeling, and review outputs.

#### Ethics statement

This study used fully simulated data and did not involve human participants, human samples, patient records, or prospective interventional research; therefore, ethics committee approval and informed consent were not required.

### Exploratory modeling

Two supervised models were attempted. Logistic regression was specified for the binary responder endpoint using dose, LDH, CRP, ctDNA fraction, AUC, and mutation status as predictors. Features were standardized before model fitting. However, the notebook was programmed to stop model fitting if the outcome lacked class variation. Random forest classification was then used to model the broader clinical-benefit endpoint with predictors including dose, age, ECOG, LDH, CRP, ctDNA fraction, AUC, Cmax, and mutation status. A hold-out split was used for exploratory evaluation, and model outputs included ROC AUC, feature importance ranking, and a classification report. This combined use of logistic regression and random forest was intentional. Logistic regression remains a strong baseline for many clinical-prediction tasks because of its interpretability and robustness, while random forest offers a flexible nonlinear alternative. Yet broader evidence suggests that more complex machine-learning methods do not guarantee meaningful performance gains without careful design, reporting, and validation (Christodoulou et al., 2019; Collins et al., 2024).

The datasets, analysis notebooks, and related study materials are openly available in the GitHub repository: https://github.com/mpetalcorin/early-phase-clinical-biomarker-pipeline.

## Results

### Simulated cohort captures a conservative early-phase disease profile

The final integrated analysis dataset contains 120 patients and 30 variables. The dose-group distribution includes 58 patients in the low-dose group, 31 in the medium-dose group, and 31 in the high-dose group. Across the full cohort, mean tumor percent change from baseline reaches 33.9%, indicating that the simulated population is dominated by progressive or nonshrinking disease. Median survival for the overall cohort is 243.8 days, with a range of 60.0 to 400.9 days. Mean baseline LDH is 259.2 U/L, mean CRP is 16.5 mg/L, mean ctDNA fraction is 0.106, mean AUC_{0-72} is 7558.0, and mean Cmax is 762.0 ng/mL. Together, these values generate a qualitatively plausible synthetic dataset for a heterogeneous advanced solid-tumor early-phase study, although the response distribution remains deliberately conservative and limits strict objective response analyses.

### Dose escalation improves clinical benefit and median survival

Dose-group summaries show a directional efficacy pattern that follows the simulation logic. Median survival increases from 229.8 days in the low-dose group to 249.8 days in the medium-dose group and 283.1 days in the high-dose group (Figure 2). The broader clinical-benefit endpoint also increases with dose, occurring in 8.6% of low-dose patients, 29.0% of medium-dose patients, and 45.2% of high-dose patients. These findings indicate that although the simulation does not generate formal RECIST-like responses, it does encode increasing disease control with increasing dose. Biomarker levels vary across dose groups without following a simple monotonic pattern. Mean LDH measures 258.9 U/L in the low-dose group, 284.1 U/L in the medium-dose group, and 234.8 U/L in the high-dose group. Mean CRP measures 14.1, 13.6, and 24.1 mg/L in the low-, medium-, and high-dose groups, respectively. This variation reflects the stochastic structure of the simulation and supports a heterogeneous biological background rather than a purely dose-driven biomarker distribution.

**Figure 2.**
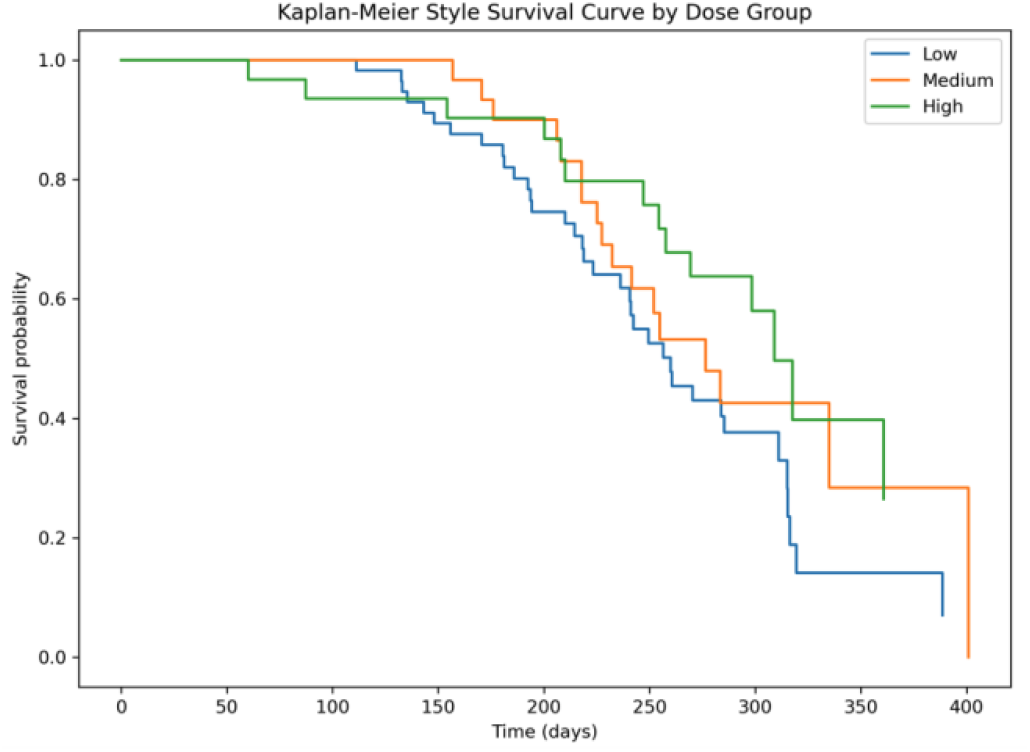
Kaplan-Meier-style survival curves by dose group. Kaplan-Meier-style plot showing survival probability over time for patients grouped into low-, medium-, and high-dose cohorts. The curves illustrate a dose-related separation in survival patterns, with the high-dose group tending to maintain higher survival probability across follow-up compared with the low-dose group.

### Strict responder classification fails because no patient reaches the RECIST-like threshold

No patient meets the prespecified binary responder threshold of at least 30% tumor reduction from baseline. As a result, the responder rate remains 0% in every dose group. This outcome makes the broader clinical-benefit endpoint more informative than the strict responder endpoint in the present dataset. The absence of formal responders shows that dose-related improvement is present, but not calibrated strongly enough to cross the conventional RECIST-like partial response threshold.

### Safety signals remain plausible without showing a monotonic dose trend

Safety outputs remain within a plausible exploratory range across dose groups (Figure 3). Grade 3+ adverse event rates are 10.3% in the low-dose group, 12.9% in the medium-dose group, and 9.7% in the high-dose group. Serious adverse event rates are 6.9%, 9.7%, and 3.2%, respectively, while discontinuation rates are 17.2%, 16.1%, and 9.7%. These data show that the synthetic safety signal does not increase monotonically with dose. Instead, the observed pattern suggests that modest sample size, random variability, and patient-level covariates contribute meaningfully to toxicity outcomes.

**Figure 3.**
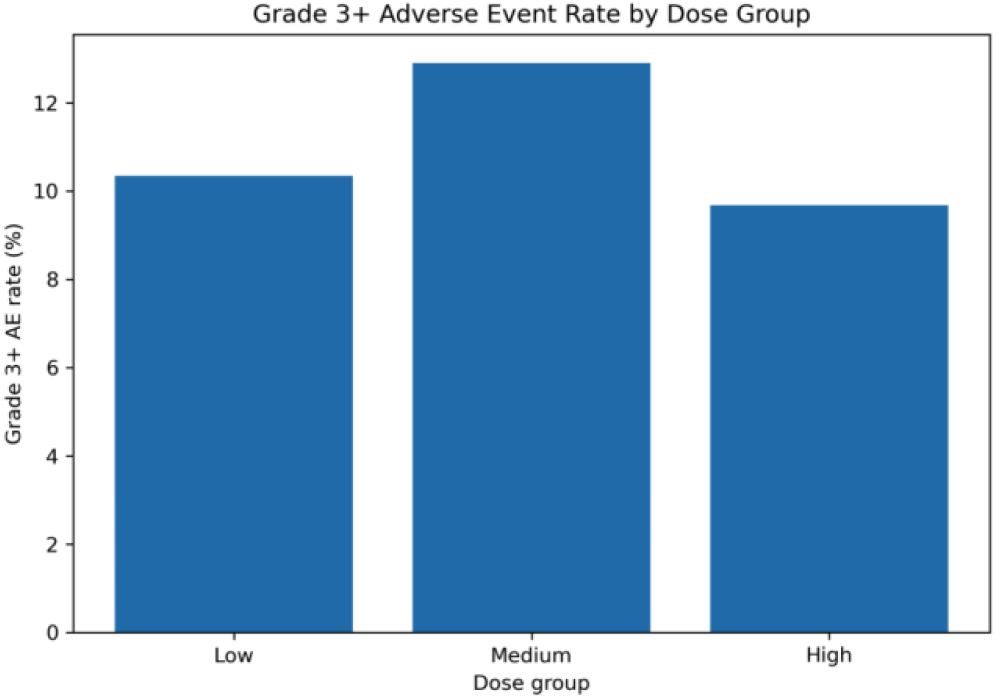
Grade 3 or higher adverse event rate by dose group. Bar chart showing the proportion of patients with grade 3 or higher adverse events in the low-, medium-, and high-dose groups. The figure summarizes severe toxicity burden across dose levels and provides an exploratory safety comparison within the simulated cohort.

### Waterfall plotting reveals heterogeneous tumor change but no objective shrinkage

The waterfall plot reveals substantial heterogeneity in tumor change across individual patients (Figure 4). Tumor percent change ranges from approximately −8.5% to +76.3%. Most bars lie above zero, and many exceed the +20% reference line, indicating progressive disease under RECIST-like logic. Only a small minority of patients show any degree of shrinkage, and none cross the −30% threshold. This figure emphasizes that the simulation produces variable tumor trajectories, but retains an overall pattern dominated by disease progression rather than objective response.

**Figure 4.**
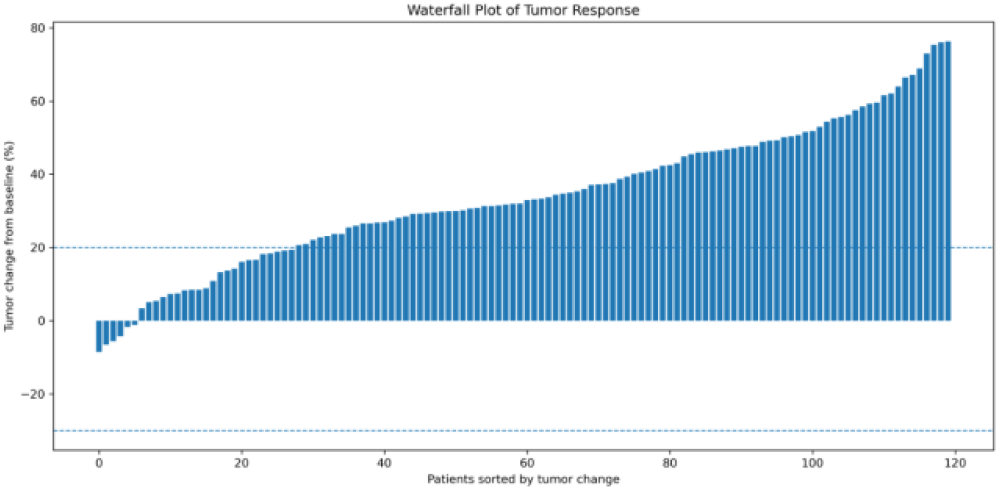
Waterfall plot of tumor response in the simulated early-phase oncology cohort. Waterfall plot showing patient-level percent change in tumor size from baseline, with patients ordered from greatest tumor reduction to greatest tumor increase. Each vertical bar represents one patient. The dashed horizontal lines at −30% and +20% indicate RECIST-like reference thresholds for partial response and progressive disease, respectively.

### Higher dose groups retain better survival over follow-up

Kaplan-Meier-style survival curves show visibly improved survival in the higher-dose groups relative to the low-dose group (Figure 2). The high-dose curve remains shifted upward across much of follow-up, consistent with the dose-group median survival values. The medium-dose group shows an intermediate pattern. These curves demonstrate that the simulation successfully encodes a dose-related benefit in time-to-event outcomes, even though it does not generate RECIST-like responders.

### High-risk biomarker status shortens survival within each dose group

Exploratory subgroup analysis further reinforces the intended biological structure. In the low-dose group, high-risk biomarker-positive patients show shorter median survival than biomarker-negative patients, 192.5 versus 240.8 days, and a higher event rate, 80.0% versus 55.8%. In the medium-dose group, biomarker-positive patients show a median survival of 206.2 days and an event rate of 100.0%, compared with 253.8 days and 46.2% in biomarker-negative patients. In the high-dose group, biomarker-positive patients again fare worse, with median survival of 267.6 versus 283.1 days and event rates of 62.5% versus 39.1%. These subgroup patterns show that adverse biomarker status continues to separate outcomes even within dose strata.

### Baseline LDH tracks with worse tumor-change trajectories

The scatter plot of baseline LDH versus tumor percent change shows a positive trend, indicating that higher LDH tends to associate with worse tumor-response trajectories (Figure 5). Although the relationship is visually modest, it aligns with the intended biological rationale that elevated LDH reflects more aggressive metabolic and disease states. The slope of the fitted line supports the direction of this effect and confirms that the biomarker simulation connects meaningfully with the tumor-response output.

**Figure 5.**
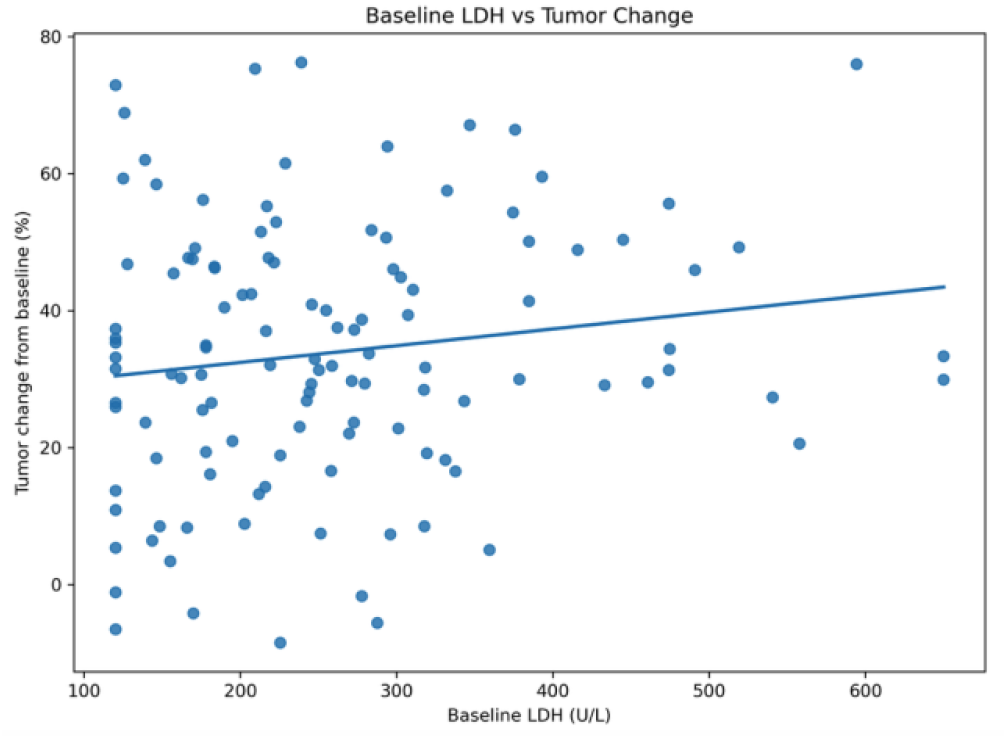
Association between baseline LDH and tumor change from baseline. Scatter plot showing the relationship between baseline lactate dehydrogenase, LDH, concentration and percent tumor change from baseline. Each point represents one patient, and the fitted line summarizes the overall linear trend.

### Inflammatory burden and ctDNA associate with tumor worsening, while exposure associates with disease control

Rank-based exploratory analyses support the visual findings. Tumor worsening shows a positive association with CRP, with Spearman rho = 0.355, and with ctDNA fraction, with Spearman rho = 0.444. In contrast, higher dose, higher AUC_{0-72}, and higher Cmax each associate with lower tumor percent change, indicating more favorable disease control, with Spearman rho values of −0.374, −0.352, and −0.365, respectively. These associations confirm that worse inflammatory and tumor-burden biomarkers track with poorer outcomes, whereas greater exposure tracks with improved early disease control.

### PK profiles separate cleanly by dose group and peak early

Mean PK profiles show the expected qualitative separation across dose groups (Figure 6). All groups display an early rise, a peak near 4 hours, and a subsequent decline. The high-dose group maintains the largest concentrations throughout the profile, followed by the medium-dose group and then the low-dose group. This pattern yields dose-dependent increases in both AUC and Cmax and provides a mechanistic bridge between administered dose and downstream clinical-benefit modeling.

**Figure 6.**
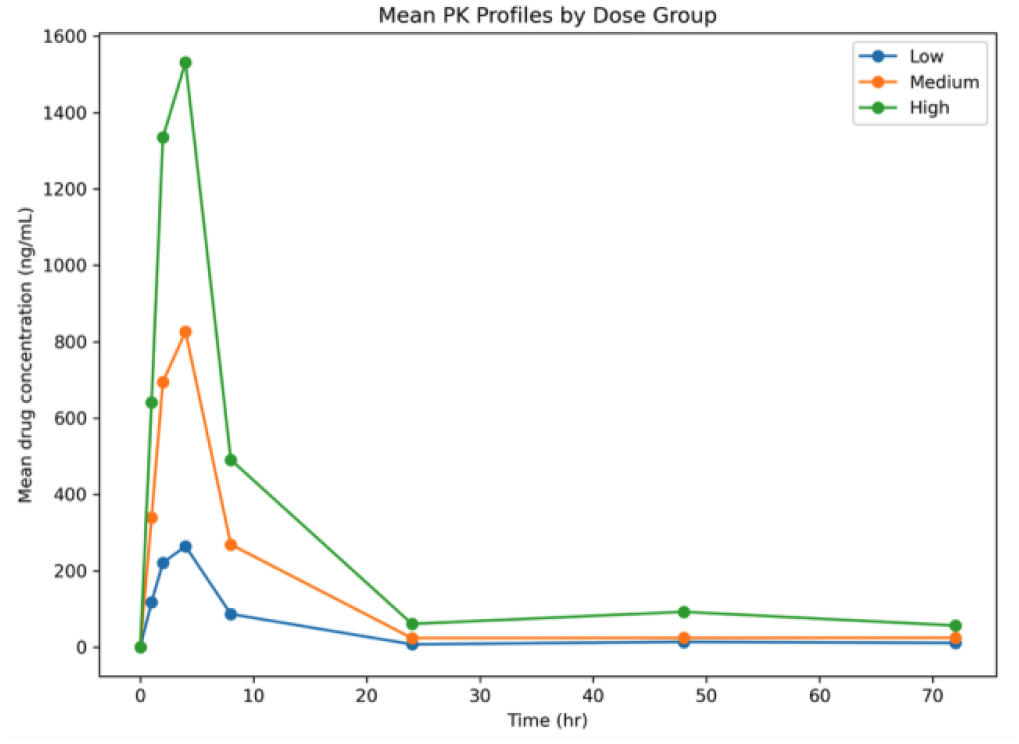
Mean pharmacokinetic profiles by dose group. Line plot showing mean drug concentration over time for the low-, medium-, and high-dose groups across scheduled pharmacokinetic sampling points. Higher dose groups show greater overall exposure and peak concentration, demonstrating the expected dose-exposure relationship built into the simulated pharmacokinetic data.

### Logistic regression fails on responder status, but random forest predicts clinical benefit

The logistic-regression model for responder status cannot be fit because the binary outcome lacks class variation. All simulated patients are labeled nonresponders under the strict threshold of at least 30% tumor reduction. This result highlights a key simulation pitfall, namely that unrealistic endpoint prevalence can render a downstream model unidentifiable.

In contrast, the random-forest model for clinical benefit fits successfully and achieves an exploratory ROC AUC of 0.845 on the hold-out set. Feature-importance ranking places Cmax, CRP, AUC_{0-72}, ctDNA fraction, LDH, age, dose, ECOG, and mutation status in descending order. In the held-out classification report, overall accuracy reaches 0.80. Performance is stronger for the majority class of no clinical benefit than for the minority class of clinical benefit, with recall for the benefit class of 0.14. This class imbalance again reflects the conservative endpoint distribution built into the simulated dataset.

### The pipeline generates coherent decision-support outputs despite conservative response prevalence

Taken together, the figures and summaries show that the workflow produces a coherent early-phase oncology dataset with internally consistent clinical, biomarker, and PK relationships (Figures 1-6). Dose escalation improves median survival and clinical benefit, adverse biomarker profiles worsen tumor and survival outcomes, and higher exposure aligns with improved disease control. At the same time, the strict responder endpoint remains absent, which limits one class of downstream modeling and underscores the importance of calibrating simulation frameworks to the intended analytical question.

## Discussion

This project shows that a compact and transparent Python workflow can generate a coherent early-phase oncology analytical environment from fully synthetic inputs. The notebook reproduces several components that are central to real early development work, including the generation of heterogeneous source data, integration of clinical, biomarker, and pharmacokinetic tables, derivation of analysis-ready variables, production of clinician-facing figures and tables, and fitting of pragmatic exploratory models. In practical oncology development, these tasks are rarely performed in isolation. Instead, pharmacokinetic exposure, biomarker behavior, safety, and early efficacy signals are increasingly interpreted together in order to inform dose selection, characterize therapeutic window, and prioritize next-step development decisions (Gibbs et al., 2010; Wang et al., 2017; Solans et al., 2020). The present workflow reflects that integrated logic and therefore serves as a useful proof-of-concept for translational programming in early oncology studies.

A major strength of the workflow is that it encodes biologically interpretable dependencies rather than generating disconnected random variables. Higher dose and higher exposure, reflected by AUC and Cmax, align with improved clinical benefit and better survival patterns, whereas higher LDH, CRP, and ctDNA align with less favorable tumor-response and time-to-event outcomes. These dependencies were intentionally chosen because they reflect recurrent translational themes in oncology. Circulating tumor DNA has become an increasingly important biomarker for disease burden assessment, treatment monitoring, and early signal detection in clinical trials, especially when serial tissue sampling is impractical (Parisi et al., 2023; Tan et al., 2026). LDH has long been associated with aggressive disease biology, metabolic stress, and worse prognosis across many solid tumors, while CRP reflects systemic inflammation and often tracks with poorer outcomes, recurrence risk, and treatment resistance (Petrelli et al., 2015; Shrotriya et al., 2015). By embedding these biologically plausible relationships into the simulation, the workflow produces outputs that are easier to interpret and more relevant to real translational decision-making.

Another important strength is the explicit creation of an integrated analysis-ready dataset. In many translational oncology settings, the hardest part of the analytical workflow is not the final model, but the intermediate data-engineering step. Clinical variables, biomarker measurements, and PK samples are usually collected at different levels of granularity, at different time points, and often under partially different conventions. In real projects, analysts must reconcile patient identifiers, align timing windows, define derivation rules, summarize longitudinal signals, and generate variables suitable for review and modeling. That practical layer is often underappreciated, even though it strongly shapes the validity and reproducibility of downstream analysis. By producing raw source files, a merged processed dataset, summary tables, patient listings, figures, and model outputs, the present workflow mirrors the real path from data assembly to exploratory interpretation. This structure also aligns with the broader emphasis on transparency promoted by TRIPOD and TRIPOD+AI, where clear reporting of variable definitions, derivation logic, analytic assumptions, and model outputs is treated as essential rather than optional (Collins et al., 2015; Collins et al., 2024).

The results also illustrate an important conceptual point about early-phase studies. In many such studies, especially those involving targeted agents or biologically selective drugs, clinically meaningful activity may appear first as disease stabilization, delayed progression, biomarker improvement, or exposure-dependent trends rather than frequent RECIST-defined objective responses. In the present simulation, no patient met the strict threshold for partial response, yet clinical benefit increased across dose groups and survival patterns shifted in the expected direction. This kind of result is not meaningless. On the contrary, it resembles the reality that early development often depends on assembling a mosaic of weak-to-moderate signals rather than waiting for a single definitive endpoint. A dose level that produces better disease control, more favorable biomarker patterns, and longer survival without a large toxicity penalty may still be informative for subsequent development, even if formal response rates remain low. In that sense, the workflow captures an important feature of translational oncology, namely that decision-support often depends on triangulating across multiple imperfect indicators.

The project further highlights both the promise and the limitations of simple machine-learning approaches in this context. The random-forest model for clinical benefit achieves an exploratory ROC AUC of 0.845 and identifies a sensible ranking of influential predictors, with exposure and biomarker measures appearing near the top. This finding is encouraging because it suggests that the synthetic dataset contains coherent signal structure and that the clinical-benefit endpoint is at least partially learnable from the derived features. At the same time, the recall for the minority class remains poor, and the logistic-regression model for strict responders cannot be fit at all because the responder endpoint has no class variation. These results provide a useful practical lesson. Performance metrics alone do not tell the full story. The usefulness of any model depends critically on how the endpoint is defined, how common that endpoint is in the dataset, and whether the data-generation process creates realistic separability between classes. This observation is consistent with the broader literature showing that complex machine-learning methods do not automatically outperform simpler regression models in clinical prediction settings, and that transparent baselines often remain indispensable (Christodoulou et al., 2019).

From a simulation-design standpoint, the absence of RECIST-like responders is the most important weakness of the current workflow. The waterfall plot shows some degree of shrinkage in a minority of patients, so the system does generate heterogeneity and a directional dose effect, but the favorable shift is not large enough to push any patient across the −30% threshold for partial response. Under RECIST 1.1, that threshold is central to the definition of objective response in solid tumors (Eisenhauer et al., 2009; Schwartz et al., 2016). Therefore, a simulated dataset intended to support responder modeling should generally produce at least a small but nonzero prevalence of such events, unless the specific aim is to represent a refractory or mainly cytostatic setting. Because the current dataset yields zero responders, any attempt to fit a model for strict objective response becomes mathematically and conceptually uninformative. This is not merely a technical inconvenience. It exposes a deeper issue in simulation work, namely that realistic endpoint prevalence is just as important as realistic distributions of baseline features.

That limitation is also scientifically useful because it reveals why simulation-based analytical projects should include calibration diagnostics, not just code execution. A workflow can run successfully and still generate data that are poorly matched to the intended modeling task. In this case, better calibration could be achieved in several ways. The favorable dose effect on tumor change could be increased, the adverse penalty attached to LDH, CRP, or ctDNA could be moderated, or a molecularly defined subgroup with enhanced sensitivity could be introduced. Any of these changes could produce a more realistic minority of objective responders and thereby permit fairer comparison between logistic regression and nonlinear machine-learning methods. More broadly, this example shows that simulation is not a one-step act of data generation, but an iterative process of design, evaluation, calibration, and redesign.

The survival component of the workflow also warrants careful interpretation. Kaplan-Meier-style curves were used because they are intuitive, familiar, and visually effective for early descriptive review. They are often the first tool that teams reach for when examining time-to-event outcomes across dose groups or biomarker-defined strata. However, Kaplan-Meier curves do not by themselves solve the inferential complexities of survival analysis. Their interpretation can become misleading when there are competing risks, strong nonproportional hazards, or informative censoring (D’Arrigo et al., 2021; Lacny et al., 2018; Gomes et al., 2024). The present workflow intentionally uses these curves descriptively rather than inferentially, which is appropriate for a proof-of-concept notebook. Still, future versions could be strengthened by adding Cox proportional hazards models, time-dependent covariate structures, restricted mean survival time analyses, or competing-risk frameworks when the synthetic design calls for them. Such additions would make the survival component more methodologically complete and more closely aligned with the range of questions encountered in oncology development.

The safety outputs also deserve comment. Grade 3+ adverse events, serious adverse events, and discontinuations remain within a plausible exploratory range, but they do not show a strictly monotonic relationship with dose. That result is realistic in one sense, because real early-phase datasets often show noisy and irregular safety patterns due to sample size, patient heterogeneity, prior treatment burden, and interacting risk factors. At the same time, it reminds the reader that the simulation does not attempt to model all biological or operational contributors to toxicity. Safety is represented in a pragmatic rather than mechanistic manner. For a pedagogic workflow, this is acceptable. For a more advanced translational simulation, however, one might wish to model cumulative toxicity, dose delays, dose reductions, exposure-toxicity relationships, and event timing more explicitly.

The project should also be interpreted within the limits of its intended use. It is not a regulatory workflow, does not implement CDISC structures, does not perform confirmatory statistical inference, and does not rely on real patient records. It is instead a methodological and educational demonstration. Its value lies in showing how a programmer or translational scientist can move from conceptually rich but heterogeneous early-development variables to a reproducible suite of outputs that support review, discussion, and refinement. In that sense, the workflow functions not only as an analysis example, but also as a communication artifact. It shows stakeholders how raw scientific data can be transformed into an interpretable package of tables, plots, and model summaries that are suitable for multidisciplinary discussion.

This is particularly relevant in early oncology development, where communication across disciplines is crucial. Clinical pharmacologists may focus on exposure, translational scientists may focus on biomarker shifts, clinicians may focus on response or tolerability, and data scientists may focus on prediction or pattern detection. A reproducible integrated workflow helps bridge those perspectives. By placing all of the relevant outputs in a traceable structure, the workflow encourages consistent interpretation and reduces the risk that different groups will work from partially incompatible summaries. This kind of integration is often more valuable in practice than a marginal gain in model complexity.

Future versions of the workflow could improve realism and utility in several directions. Longitudinal biomarker modeling would allow the study of within-patient changes over time rather than relying only on baseline biomarker burden. Formal exposure-response regression could quantify the relationship between AUC or Cmax and specific efficacy or toxicity endpoints rather than treating exposure mainly as a descriptive feature. Repeated-measures tumor trajectories could replace the current single time-point tumor assessment and thereby better mimic serial radiographic review in oncology trials. Dose interruptions, reductions, and treatment discontinuations could be modeled as dynamic rather than static outcomes. Biomarker-defined treatment-effect heterogeneity could be introduced to emulate targeted therapy settings more closely. Disease-specific external parameterization from phase I trial literature could also improve realism and allow the workflow to represent specific tumor types rather than a generic advanced solid-tumor population.

From a software and portfolio perspective, the repository could also be expanded in ways that enhance professional usefulness. CDISC-like variable naming and metadata could be added for training purposes. Automated report generation could produce a regular review package for internal study meetings. Interactive dashboards could support exploratory filtering by dose group, biomarker status, or outcome class. Calibration modules could formally assess whether simulated event rates, response distributions, and biomarker ranges are suitable for the intended downstream models. Together, such additions would transform the workflow from a good proof-of-concept into a more mature demonstration of early development programming and translational analytics.

In summary, the present project succeeds in its main purpose. It creates a reproducible and interpretable synthetic oncology dataset, integrates multiple data domains into an analysis-ready structure, and generates outputs that resemble the kinds of exploratory summaries used in early development review. Its strongest features are transparency, internal biological coherence, and practical relevance to translational workflow design. Its main weakness, the absence of objective responders, is also its most instructive lesson because it exposes the importance of simulation calibration for downstream analytical validity. As a result, the workflow is useful not only as a demonstration of coding and data integration, but also as a demonstration of how analytical design choices shape scientific interpretation.

## Conclusion

A reproducible computational pipeline successfully simulates and analyzes an early-phase oncology trial by integrating clinical variables, translational biomarkers, and pharmacokinetic measurements into a single analytical workflow. The pipeline generates an analysis-ready dataset, traceable summary tables, standard oncology visualizations, and exploratory predictive models, thereby demonstrating how heterogeneous early development data can be transformed into interpretable decision-support outputs. The synthetic results preserve the intended biological and translational structure, with higher biomarker burden aligning with poorer tumor and survival outcomes, and greater drug exposure aligning with improved clinical benefit.

At the same time, the project identifies an important calibration limitation. The strict responder endpoint shows zero prevalence, which prevents logistic-regression modeling of objective response. This does not weaken the overall value of the workflow. Rather, it highlights a central principle in translational analytics, namely that reproducibility by itself is insufficient if endpoint definitions, event frequencies, and simulated variable distributions are not well matched to the intended downstream question. In both simulated and real-world oncology pipelines, analytical usefulness depends not only on code that runs reliably, but also on data structures that are scientifically and statistically fit for purpose.

Taken together, this proof-of-concept provides a practical and extensible foundation for early development programming, exploratory biomarker analytics, and oncology-focused data-science training. With further calibration and expansion, the framework can support more realistic exposure-response analyses, longitudinal modeling, and translational workflow development for early-phase cancer research.

## Data Availability

All data generated in this study are simulated and are available in the GitHub repository for this project, together with the analysis notebooks and related study materials: https://github.com/mpetalcorin/early-phase-clinical-biomarker-pipeline

https://github.com/mpetalcorin/early-phase-clinical-biomarker-pipeline

